# Modelling the impact of household size distribution on the transmission dynamics of COVID-19

**DOI:** 10.1101/2021.01.12.21249707

**Authors:** Pengyu Liu, Lisa McQuarrie, Yexuan Song, Caroline Colijn

## Abstract

Under the implementation of non-pharmaceutical interventions such as social distancing and lockdowns, household transmission has been shown to be significant for COVID-19, posing challenges for reducing incidence in settings where people are asked to self-isolate at home and to spend increasing amounts of time at home due to distancing measures. Accordingly, characteristics of households in a region have been shown to relate to transmission heterogeneity of the virus. We introduce a stochastic epidemiological model to examine the impact of the household size distribution in a region on the transmission dynamics. We choose parameters to reflect incidence in two health regions of the Greater Vancouver area in British Columbia and simulate the impact of distancing measures on transmission, with household size distribution the only different parameter between simulations for the two regions. Our result suggests that the dissimilarity in household size distribution alone can cause significant differences in incidence of the two regions, and the distributions drive distinct dynamics that match reported cases. Furthermore, our model suggests that offering individuals a place to isolate outside their household can speed the decline in cases, and does so more effectively where there are more larger households.

## 1. Introduction

The novel coronavirus disease (COVID-19) caused by severe acute respiratory syndrome coronavirus 2 (SARS-CoV-2) has created a global pandemic with over 50 million confirmed cases and more than one million deaths as of November 2020. In the absence of an effective cure and vaccine, various non-pharmaceutical interventions (NPIs), including hand hygiene, face masks, quarantine, isolation, contact tracing, and social distancing, have been the primary practices for reducing the spread of the highly transmissible respiratory pathogen. Amid these interventions, stay-at-home policies and quarantine or isolation strategies may alter social interactions and hence the transmission dynamics of the virus, especially the transmission probabilities within and outside households [1, 18].

It has been shown that the general secondary attack rate of COVID-19 to individuals within households is higher than that of severe acute respiratory syndrome (SARS) and Middle East respiratory syndrome (MERS) [7]. With higher contact rates within households under stay-at-home policies and strict lockdowns, investigating the connections between household characteristics and transmission dynamics of the virus could provide insights for designing interventions to prevent infection. A number of studies have found heterogeneity in the prevalence, hospitalization, and mortality of COVID-19 related to demographic and ethnic differences among households and household size or household density. The findings indicate that individuals from ethnic minority backgrounds, especially south Asian and black individuals, are of higher risk related to COVID-19 [9, 17, 19], and household size may be associated with the risk of infection after implementing social distancing or stay-at-home policies [8]. A geospatial analysis has investigated the connections between socioeconomic factors in households and the prevalence of the disease, and indicates that lower educational attainment and higher household occupancy are among significant risk factors of infection [3]. An analysis based on detailed patient and contact tracing data has revealed that the average risk of transmission is positively associated with the closeness of social interactions, with highest risk within households, especially during lockdowns [16]. Other studies that consider household size show that controlling transmission within households is key to successfully bringing cases into a decline [11], and that small households are preferable for curbing an outbreak during a lockdown [14]. However, how different distributions of household size would affect transmission dynamics of the virus and the effectiveness of public health policies remains unknown.

The Greater Vancouver area incorporates two regional health authorities, dividing the metropolitan area into the Fraser Health region (FH) and the Vancouver Coastal Health region (VCH). The numbers of COVID-19 cases in the two regions differ considerably, with approximately 18,000 total cases in FH and 8,000 cases in VCH as of November 2020. The household size distributions of the two regions are also different. There are 1,695,150 individuals living in 631,135 private households in FH and 1,135,295 individuals living in 493,515 private households in VCH, according to Statistics Canada 2016 Census [15].

To investigate the extent to which household size distribution affects the spread of the virus, we develop an individual-based Markov-chain SEIR model. We inform the model with data on the household size distribution in FH and VCH and with incident case data. We then analyze the impacts of household size distribution on the incidence of COVID-19 in FH and VCH, the probability of remaining uninfected for individuals living in households of different sizes, and the effectiveness of various isolation strategies.

## 2. Methods

### 2.1. Data

We obtained the household size distributions in the Fraser Health region (FH) and the Vancouver Coastal Health region (VCH) based on Statistics Canada 2016 Census [15], which includes 631,135 and 493,515 private households in FH and VCH respectively. The data contain the size of private households in British Columbia and census subdivisions of British Columbia, listing the number of households with one to seven individuals and with at least eight individuals in each subdivision. We collect the data for all census subdivisions in Fraser Health and Vancouver Coastal health regions and compute the total numbers and proportion of households of each size. COVID-19 data, including the daily incident cases in the Greater Vancouver area, are publicly available at BCCDC website [2], containing information of the dates and health regions of reported cases. We use the number of incident cases from each day between March 3, 2020 to December 3, 2020 in both FH and VCH.

### 2.2. Model description

We introduce an individual-based Markov-chain susceptible-exposed-infectious-recovered (SEIR) model, which describes the time dynamics of susceptible (*S*), exposed (*E*), infectious (*I*), and recovered or deceased (*R*) individuals. For the sake of simplicity, we assume that households with at least eight individuals contain exactly eight individuals, and each simulated individual resides in a household of size one to eight individuals. The household size distribution in a simulation is a discrete probability distribution indicating the number of households of each size in a region. We also assume each individual spends a fraction *δ* of 24 hours inside their household every day, and while an individual is in the community (outside the household), we assume the individual encountered *ρ* other individuals per day, not including the individuals in the same household. We suppose the transmission rate in the community per individual per day *β*_1_ is lower than the transmission rate within a household per individual per day *β*_2_, and that recovered individuals are immune to the virus.

We model transmission over *n* days in a region with *N* individuals. Let Ω = {*S, E, I, R*}^*N*^ be the sample space, and 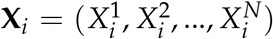 be an *N*-dimensional multivariate random variable representing the compartment each individual is in on day *i*. Specifically, 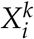 denotes the compartment individual *k* is in on day *i*. We define a discrete-time Markov chain **X**_1_, **X**_2_, …, **X**_*n*_ in the following way.

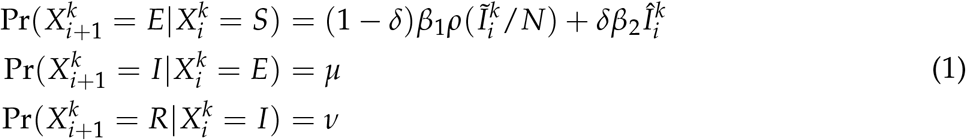

In this model, each individual may contract the virus in two ways: from the community or within the household. The symbol 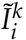 denotes the number of individuals in compartment *I* on day *i* who are not from the same household as individual *k*, and the symbol 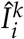 denotes the number of individuals in compartment *I* on day *i* who are from the same household as individual *k*. The top schematic diagram in Figure 1 provides a visual representation of the transmission dynamics governed by the equations (1). Note that the key parameter in this model, the household size distribution, is implicit in the formulas, as it affects the variables 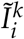 and 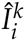.

**Figure 1:**
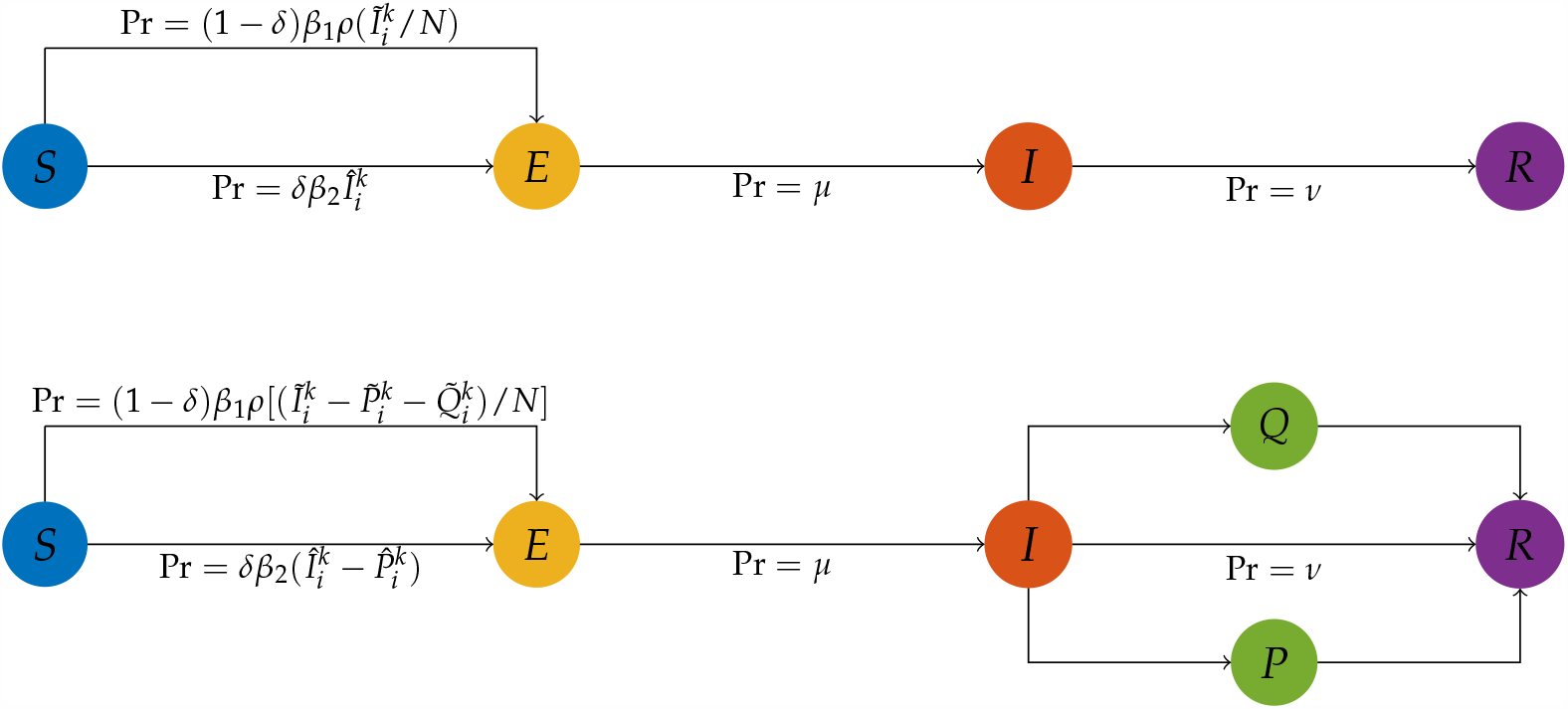
Top: the schematic diagram of the basic epidemiological model. Bottom: the schematic diagram of the adjusted epidemiological model with an isolation state, where *P* is the compartment of individuals practicing separated isolation and *Q* is the compartment of individuals practicing home isolation. The probabilities displayed in both diagrams are for each simulated individual.

### 2.3. Scenarios

We study two scenarios. The first corresponds to the reported cases in the Fraser Health (FH) and Vancouver Coastal Health (VCH) regions and investigates the potential impact of household size distribution on the incidence. The second is a hypothetical scenario examining the effectiveness of various isolation strategies under different household size distributions. For all simulations regarding the two major scenarios, we assign identical values to the universal parameters listed in Table 1. For both scenarios, we set the number of simulated individuals to *N* = 500, 000, and assume that the sample size will not affect the transmission dynamics when the number of immune individuals is less than 1% of the simulated population.

**Table 1:** List of universal parameters and their values assigned for all scenarios analyzed in this article.

| Parameter | Definition | Value |
| --- | --- | --- |
| $\beta_1$ | Transmission rate in the community per individual per day | 0.011 |
| $\beta_2$ | Transmission rate within households per individual per day | 0.09 |
| $\mu$ | Rate from compartment E to I per individual per day | 0.15 |
| $\nu$ | Rate from compartment I to R per individual per day | 0.071 |
| $\rho$ | Number of individuals encountered in the community per individual per day | 20 |

The first scenario has an initial date of February 11, 2020. We assume on the initial date, there exist 10 individuals in compartment *I* (selected uniformly at random from all simulated individuals in compartment *S*), 50 individuals in compartment *E* (selected uniformly at random from all simulated individuals in compartment *S* after selecting the individuals in compartment *I*), and no individual in compartment *R*. The length of a simulation for this scenario is 300 days, with December 7, 2020 as the last day. To model different phases of implementing stay-at-home policies and reopening, the parameter *δ* in this scenario takes on different values throughout a simulation. From the initial day to day 40 (March 21, 2020), *δ* = 0.625 representing the baseline case when each individual approximately spends 15 hours at home and 9 hours in the community on average; see Table 2 for detailed values.

**Table 2:** The values of the parameter *δ* throughout a simulation for the first scenario.

| Date | Value of $\delta$ |
| --- | --- |
| Day 1 - 40 (March 21, 2020) | 0.625 |
| Day 40 - 47 | 0.625 to 0.925 (linearly) |
| Day 47 - 140 (June 29, 2020) | 0.925 |
| Day 140 - 147 | 0.925 to 0.675 (linearly) |
| Day 147 - 210 (September 7, 2020) | 0.675 |
| Day 210 - 217 | 0.675 to 0.875 (linearly) |
| Day 217 - 240 (October 7, 2020) | 0.875 |
| Day 240 - 247 | 0.875 to 0.675 (linearly) |
| Day 247 - 300 (December 7, 2020) | 0.675 |

The second scenario is hypothetical and concerns isolation strategies. We adopt the same assumptions and initial conditions as in the first scenario.Moreover, we assign each individual a preference regarding how they would practice isolation if they are in compartment *I* when isolation is recommended. The possible preferences for an individual are: not practicing isolation, practicing isolation at home, and practicing isolation at a separated place. We assume that individuals who prefer to not practice isolation can infect any other individual in compartment *S*, individuals who prefer to practice isolation at home can infect only individuals in compartment *S* in the same household, and individuals who prefer to isolate at a separated place cannot infect any individual in compartment *S*. To model different isolation strategies, we modified the model according to (2), where 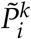 and 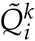 denote the number of individuals who are not in the same household as individual *k*, and who are practicing separated isolation and practicing home isolation on day *i* respectively. 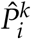 denotes the number of individuals who are in the same household as individual *k* and who are practicing separated isolation on day *i*. For a visual representation of this model, see the bottom schematic diagram in Figure 1.

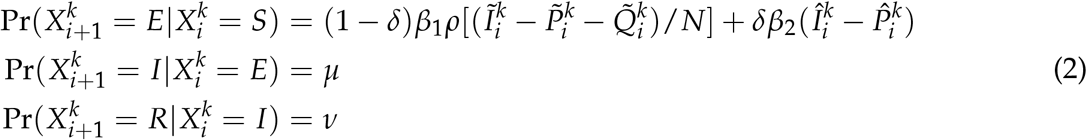

In this scenario, the parameter *δ* = 0.675 remains constant for all simulated days and the length of a simulation is 200 days. From the initial day to day 50, infected individuals are not recommended to isolate, meaning no individual practices any type of isolation. Starting from day 51 to the end of a simulation, the simulated individuals practice isolation with respect to their preferences.

To compare the effectiveness of different isolation strategies in regions with different household size distributions, we designed four isolation scenarios listed in Table 3. The only parameters that vary between the isolation scenarios are the household size distribution (between FH and VCH) and the distribution of the isolation preferences over the simulated individuals. We keep all other parameter values in each of the simulations identical for all isolation scenarios. The isolation scenario FH-H (home isolation) uses the household size distribution in FH. 55% of the simulated individuals would practice isolation at home when they are in compartment *I* while the other 45% simulated individuals would not practice isolation. Similarly, the isolation scenario FH-S (separated isolation) also uses the household size distribution in FH, with the difference that 55% of the simulated individuals would practice isolation at a separated place and the remaining 45% would not practice isolation. The distribution of isolation preferences over the simulated individuals in isolation scenarios VCH-H and VCH-S is the same as in scenarios FH-H and FH-S, respectively, but these isolation scenarios are with the household size distribution in VCH.

**Table 3:**
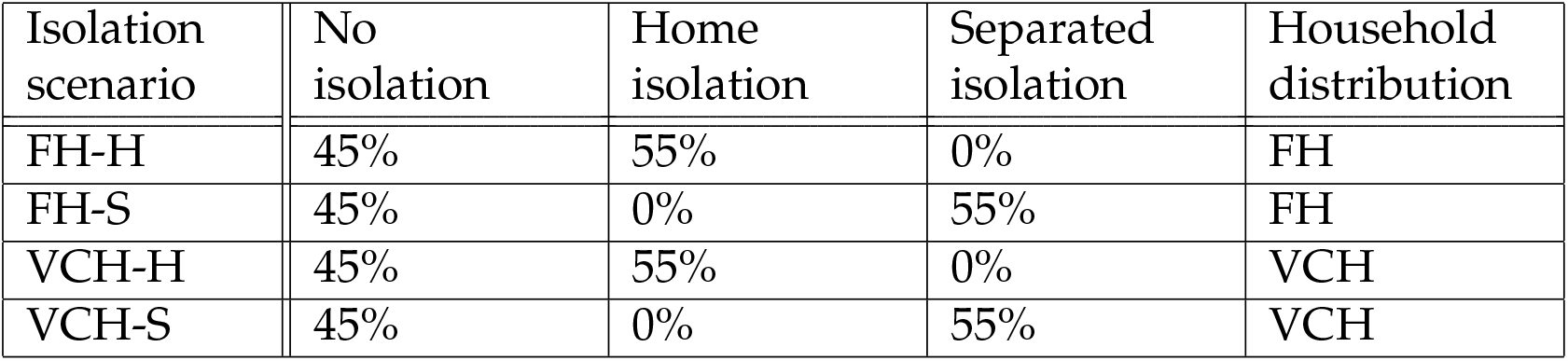
List of isolation scenarios, corresponding isolation preferences and household size distribution in use.

| Isolation scenario | No isolation | Home isolation | Separated isolation | Household distribution |
| --- | --- | --- | --- | --- |
| FH-H | 45% | 55% | 0% | FH |
| FH-S | 45% | 0% | 55% | FH |
| VCH-H | 45% | 55% | 0% | VCH |
| VCH-S | 45% | 0% | 55% | VCH |

### 2.4. Probability of remaining uninfected

We use survival analysis techniques to analyse the probability of an individual becoming infected on each simulated day. We apply a Kaplan-Meier estimator [5] to estimate the probability of remaining not infected for individuals in households of different sizes. Note that the event of interest here is the infection of an individual, so “survival” means remaining uninfected. Let *N*^*k*^ be the number of individuals in households of size *k*, and 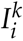 be the number of incident cases from households of size *k*, on day *i*. The survival function *L*^*k*^ (*t*), indicating the probability of remaining uninfected for individuals from households of size *k* on day *t*, is defined by the standard formula: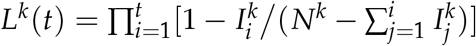.

## 3. Results

### 3.1. Distribution comparison

The household size distributions in the two health regions are different; the average household size is 2.68 in FH and is 2.31 in VCH. There are more large households and fewer single-individual households in FH; see Figure 2. We compare the distributions by Pearson’s chi-squared test [13], which rejects the null hypothesis that the household sizes in FH and VCH originate from populations with the same distribution with a *p*-value 2.2e-16. This indicates that there are differences between the household size distributions in the two health regions.

**Figure 2:**
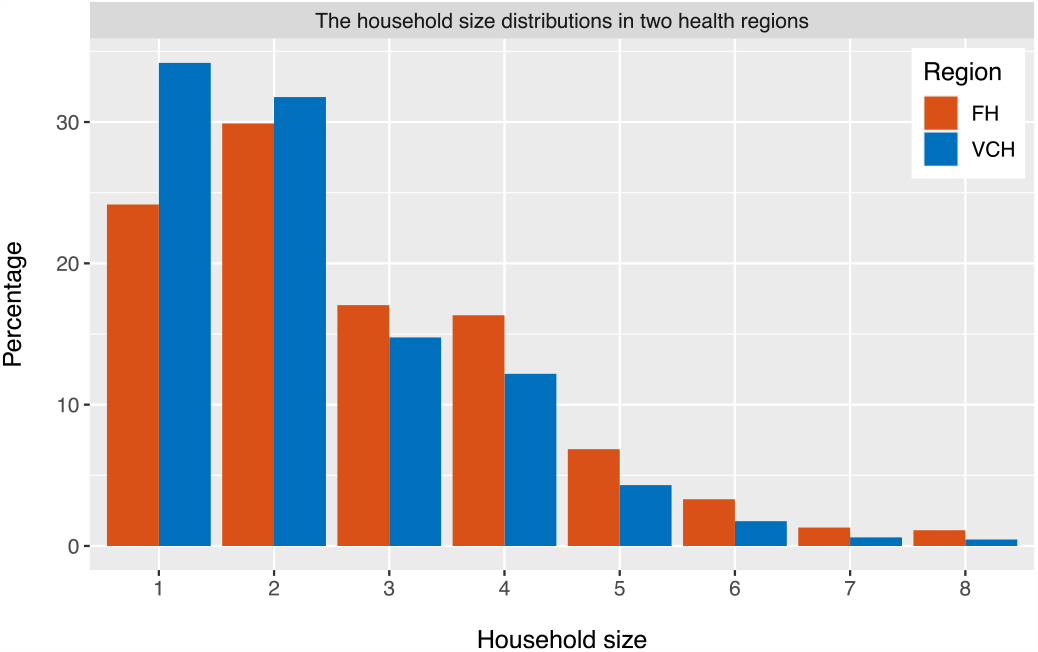
The household size distributions in the Fraser Health region and the Vancouver Coastal Health region.

### 3.2. Impact on incidence

We apply the model to analyze the impact of household size distribution on the incidence of COVID-19 in FH and VCH. We set the simulations for FH and VCH with the same initial values and parameters described in Section 2.3, except for the household size distribution, which is initialized to match the observed household size distribution in each health region. We repeat the simulation for each health region 100 times; Figure 3 displays the results. The top two panels show the number of incident cases from simulations and reported data. Note that the simulation results match the reported cases and the only parameter that differs between simulations for the left and right panels is the household size distribution. While reported cases likely do not represent all cases, for simplicity we assume a constant ascertainment fraction. These results indicate that the difference between the household size distributions in FH and VCH can lead to a substantial difference in COVID-19 incidence, suggesting that the household size distribution may be a factor causing the heterogeneity in the number of COVID-19 cases in FH and VCH.

**Figure 3:**
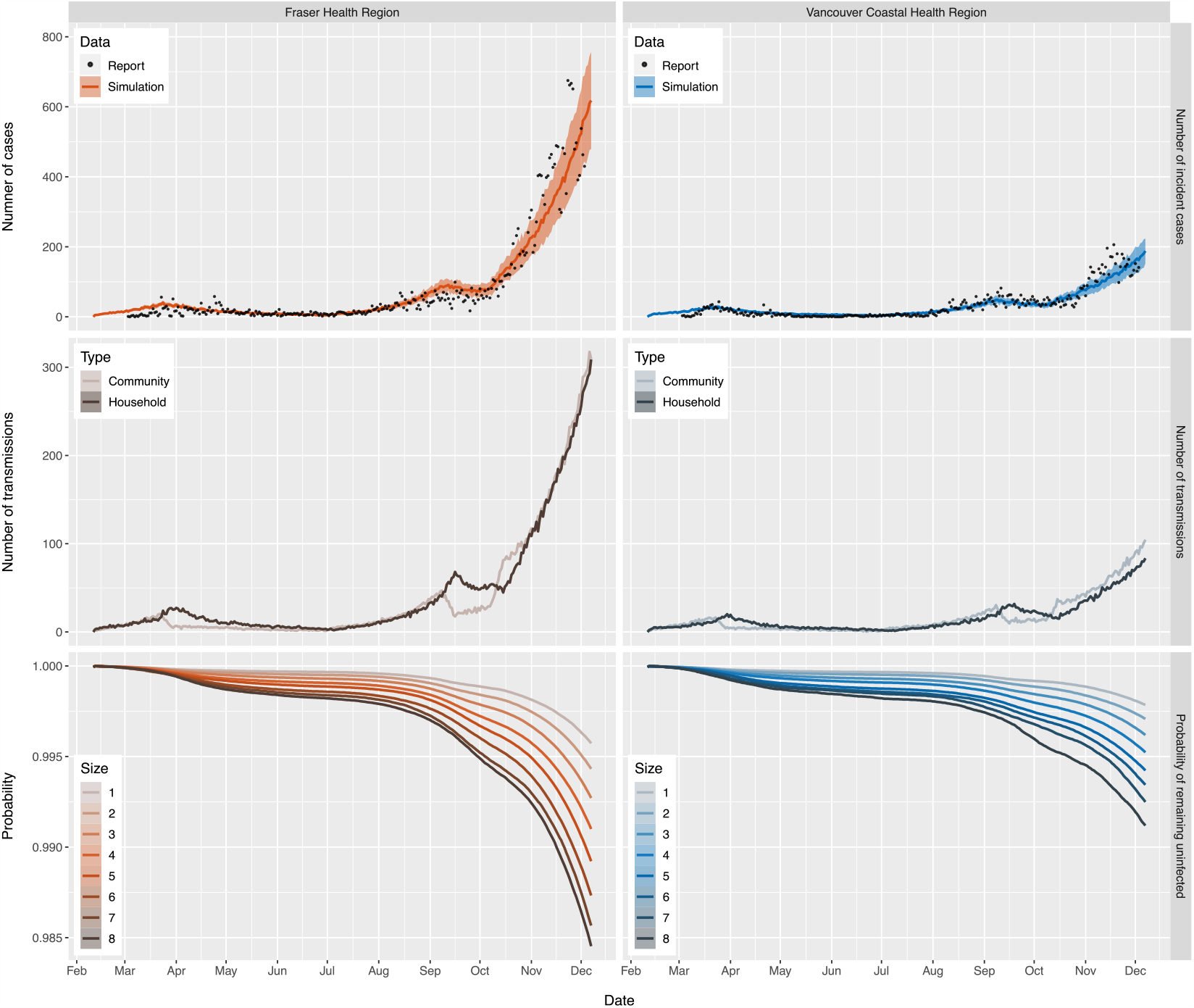
Top: the number of incident cases from simulation (curves with 95% confidence interval bands) and reported data (points) in the two regions. Middle: the number of community and household transmissions from simulations in the two regions. Bottom: the survival curves for individuals in households of different sizes from simulations in the two regions. All curves in the figure reflect mean values over 100 runs for each of the simulations which only differ in household size distribution. Model parameters are the same in the two regions, except for the household size distribution.

We also plot the number of transmissions in the community and within households (middle two panels of Figure 3). The results show that under the settings described in Section 2.3, the number of transmissions in the community and within households are similar. When stay-at-home policies are implemented, the number of community transmissions decreases but the number of household transmissions keeps increasing for several days.

### 3.3. Probability of remaining uninfected

The bottom panels of Figure 3 show the mean probability of remaining uninfected up to day *n*, over 100 runs of the simulations, for both health regions. Note that the probabilities in these plots depend on the total population size in the model (here 500,000 individuals). Without knowledge of the ascertainment fraction or the true number of individuals at the beginning of the simulation, it is not possible to relate the model’s probability of remaining uninfected to the true prevalence.

We find that individuals living in larger households have lower probability of remaining un-infected, and for each household size, the individuals living in FH have lower probability of remaining uninfected than individuals living in VCH, especially near the end of the simulations, due to the difference in prevalence of the two health regions. Moreover, under stay-at-home policies and social distancing measures, the probability of remaining uninfected for individuals living in large households decreases more substantially than for individuals living in small households.

### 3.4. Isolation effectiveness

Figure 4 shows the differences in incidence under different isolation strategies, based on 100 runs of the simulations for the second scenario described in Section 2.3. The top panel of Figure 4 shows the number of active cases in each of the four isolation scenarios. The results suggest that a proportion of individuals isolating at a separated place can result in more rapid decreases in cases than the same proportion of individuals isolating at home. Interestingly, with the same settings, 55% of the simulated individuals isolating at home can bring the cases into a decline under the household size distribution in VCH, while the number of cases continues to increase at a moderate rate under the household size distribution in FH. Comparing the bottom panels of Figure 4 indicates home isolation poses a lower probability of remaining uninfected than separated isolation for individuals in households of all sizes including individuals living by themselves. Moreover, 55% of individuals isolating at home under our settings would reduce the growth of both community and household transmissions, though it makes household transmission more prominent than community transmission; see the middle panels of Figure 4.

**Figure 4:**
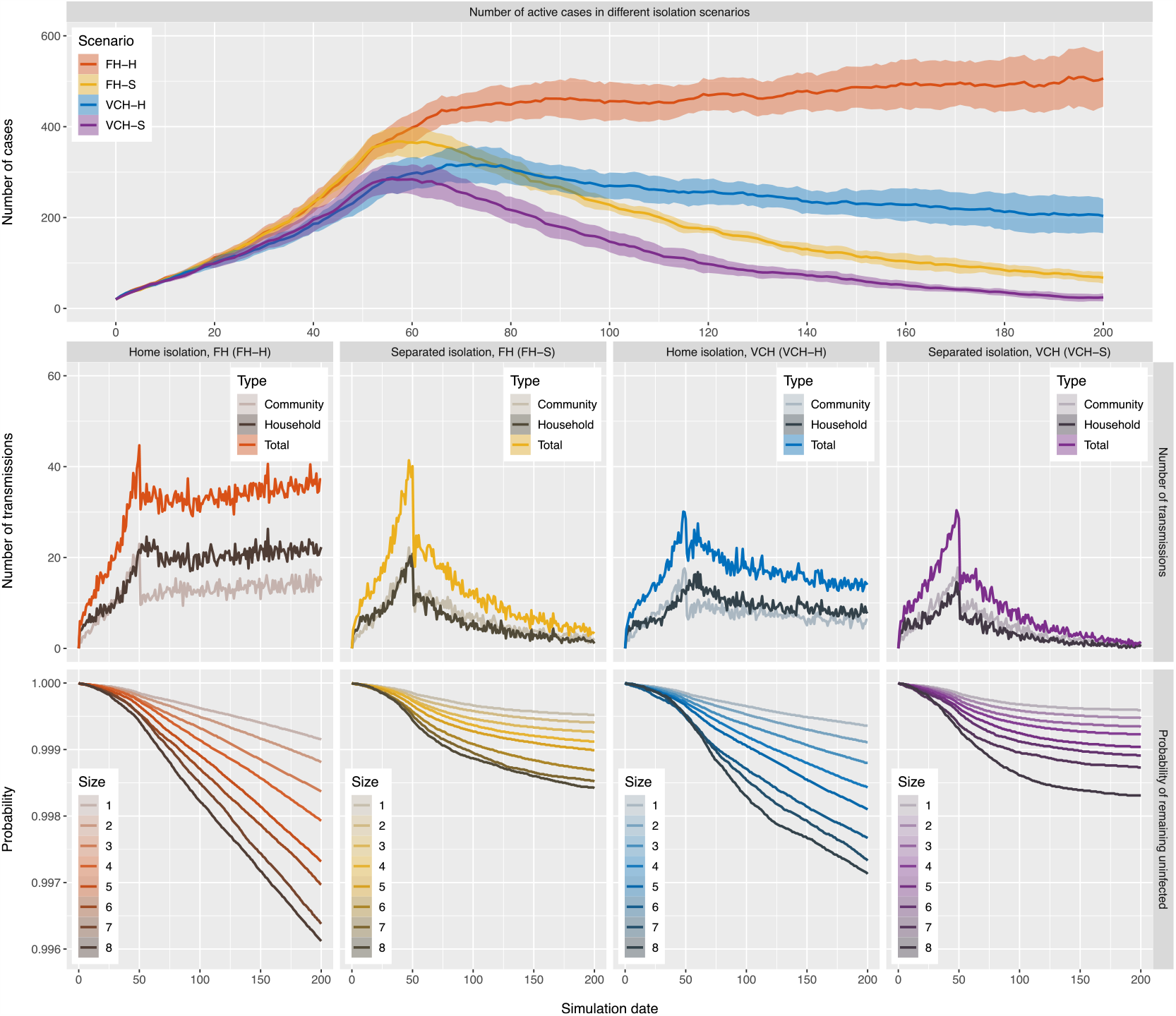
Results of simulations for isolation scenarios where 55% of the simulated individuals practice home or separated isolation. Top: the number of active cases with 95% confidence interval bands in each of the isolation scenarios. Middle: the number of transmissions of different types in each of the isolation scenarios. Bottom: the survival curves for individuals in households of different sizes in each of the isolation scenarios. All curves in the figure reflect mean values over 100 runs of the simulation for each of the isolation scenarios and the settings only differ in household size distribution and the individuals’ isolation preferences. The curves are organized so that warm colors (red and yellow) represent scenarios in FH and cold colors (blue and purple) represent scenarios in VCH.

Unless there is such widespread testing in place that individuals know they are infectious very early in their infection, an individual who becomes infectious would not practice isolation immediately, but would begin after a period of time when the individual receives a positive test or develops symptoms. We add this period of time to our simulations for isolation scenarios where 55% of individuals practicing home or separated isolation. Furthermore, we introduce an additional isolation scenario FH-HS, where home isolated individuals who live in households with four or more individuals are offered and would accept a separated place for isolation in one or two days after they start practicing home isolation. Supplementary Figure 1 shows the results. We find that home isolation after this delay is naturally less effective compared to our hypothetical experiment, and it is even more so for separated isolation in the two regions. However, it is still the case that separated isolation can result in more rapid decreases in cases than isolating individuals at home. Moreover, offering a separated isolation place for individuals who live in households with four or more individuals when 55% of the individuals practice home isolation under the household size distribution in FH can bring the cases into a decline and increase the probability of remaining uninfected for all individuals.

We also alter the proportion of simulated individuals practicing either home or separated isolation. Supplementary Figures 2 and 3 display the results for the analogous four scenarios with 75% and 25%, respectively, of the simulated individuals practicing either home or separated isolation and the rest of the individuals not practicing isolation. The results suggest that home isolation can bring cases into a decline if 75% are able to practice isolation, under either the household size distribution in FH or VCH, though in this case individuals living in larger household are of lower probability of remaining uninfected compared with separated isolation. Conversely, if not enough individuals practice isolation (here only 25%), even though some individuals practice the strict separated isolation, the intervention is insufficient to result in declining cases; see Supplmentary Figure 3.

## 4. Discussion

We have developed a stochastic model and utilized it to investigate the impacts of household size distribution and home versus separated isolation on the incidence of COVID-19. The model has been designed to be as simple as possible, with only the essential components to discover how the distributions of household size would affect transmission dynamics. Our model does not simulate the entire population of the health regions, limiting our ability to compare the absolute probability of infection. Our model also does not include an explicit simulation of contacts within and between schools, retail and social settings and workplaces, or finer geographic variation within Fraser Health and Vancouver Coastal Health regions, and indeed the data to support modelling of these complex contact structures at a high level of temporal resolution is generally not available.

We have found that under parameters reflecting COVID-19 transmission in British Columbia, the difference in household size distribution alone can account for the distinct transmission dynamics in the two health regions we have studied. We also find that in the context of directives to stay home, and to self-isolate at home if ill, an individual’s household size has a high impact on their probability of remaining uninfected. These results suggest that the household size distribution may be a key factor of transmission heterogeneity for COVID-19. Our results also show that an isolation strategy can be successful under one distribution of household size at controlling the spread of the virus but less effective under a different household size distribution, indicating that uniform policies for regions with different demographic characteristics may not be optimal. Jurisdictions with many larger households would benefit more from policies offering self-isolation at a separated place than jurisdictions with predominantly smaller households. Furthermore, at rates of transmission that are comparable to those in the Greater Vancouver area, which are likely relatively near the epidemic threshold at the time of writing, this difference could even be enough to bring COVID-19 cases into a decline.

There are a number of sources of disparity and inequity that have been found to be connected to COVID-19 risk, including the physical size of households (and therefore the density of contact), occupation [6], age [4, 7], ethnicity [9, 19], income [3, 12], and comorbidities. These intersect: larger households may have several members who are essential workers who must work outside the home, lower-income employment is less likely to allow working from home [10] and households with more members may also be more crowded. The intersection of these inequalities lends further urgency to the need to develop targeted support, including offering a separate place to isolate.

## Data Availability

Household data from Statistic Canada 2016 Census are available at Statistics Canada website. COVID-19 case data in British Columbia are available at BCCDC website.

https://www12.statcan.gc.ca/census-recensement/2016/dp-pd/index-eng.cfm

http://www.bccdc.ca/health-info/diseases-conditions/covid-19/data

## Acknowledgments

This work was supported by the grant of the Federal Government of Canada’s Canada 150 Research Chair program to Prof. C. Colijn and by Genome BC grant COV142. We thank Statistics Canada for providing the household size distribution data.

## Supplementary Material

We adjust the simulations for the second scenario defined in Section 2.3 so that simulated individuals would practice home or separated isolation in one to three days (instead of immediately) after they become infectious, which simulates the period of testing or the period prior to symptom onset. The period is chosen normally at random with mean value two days and standard deviation one day. Similarly, 55% of the simulated individuals would practice home or separated isolation in these isolation scenarios, while 45% of individuals would not practice isolation. Moreover, we introduce another isolation scenario, FH-HS (home then separated isolation), for the adjusted simulations. This isolation scenario uses the household size distribution in FH and 55% of the simulated individuals would practice home isolation in one to three days after they become infectious. Additionally, the home isolated individuals who live in households with four or more individuals are offered and would accept a separated place for isolation in one or two days (uniformly chosen at random) after they start practicing home isolation. Supplementary Figure 1 displays the result of the adjusted simulations

We also reproduce the same simulations as discussed in Section 3.4 but with altered proportions of simulated individuals practicing home or separated isolation. Supplementary Figure 2 displays the results for the four scenarios with 75% of the simulated individuals practicing either home or separated isolation and the other 25% of the individuals not practicing isolation. Supplementary Figure 3 displays the results for the four scenarios with only 25% of the simulated individuals practicing either home or separated isolation and the other 75% of the individuals not practicing isolation.

**Supplementary Figure 1:**
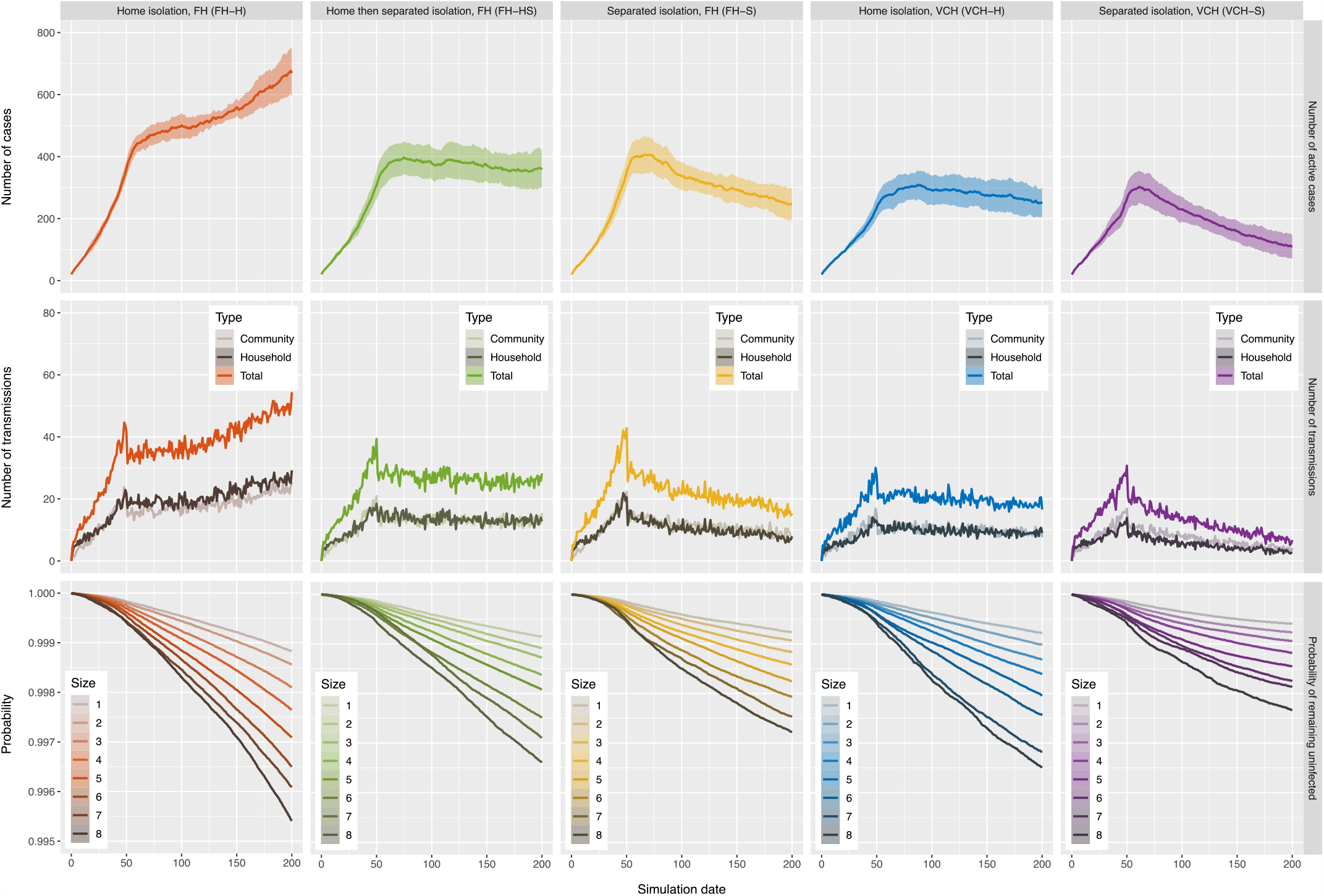
Results of adjusted simulations for isolation scenarios where 55% of the simulated individuals practice home or separated isolation after a period of one to three days. Top: the number of active cases with 95% confidence interval bands in each of the isolation scenarios. Middle: the number of transmissions of different types in each of the isolation scenarios. Bottom: the survival curves for individuals in households of different sizes in each of the isolation scenarios. All curves in the figure reflect mean values over 100 runs of the simulation for each of the isolation scenarios.

**Supplementary Figure 2:**
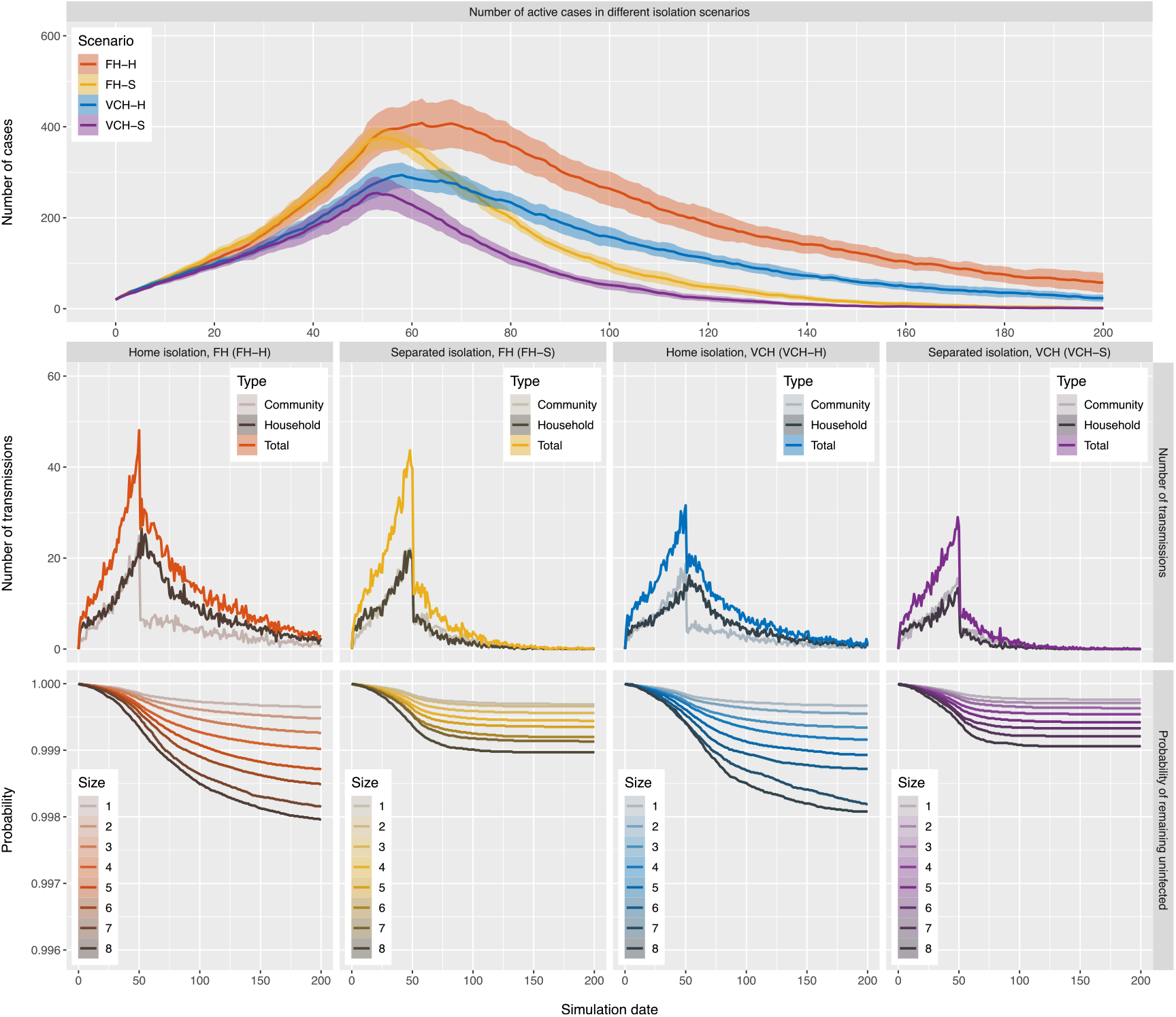
Results of simulations for isolation scenarios where 75% of the simulated individuals practice home or separated isolation. Top: the number of active cases with 95% confidence interval bands in each of the isolation scenarios. Middle: the number of transmissions of different types in each of the isolation scenarios. Bottom: the survival curves for individuals in households of different sizes in each of the isolation scenarios. All curves in the figure reflect mean values over 100 runs of the simulation for each of the isolation scenarios and the settings only differ in household size distribution and the individuals’ isolation preferences.

**Supplementary Figure 3:**
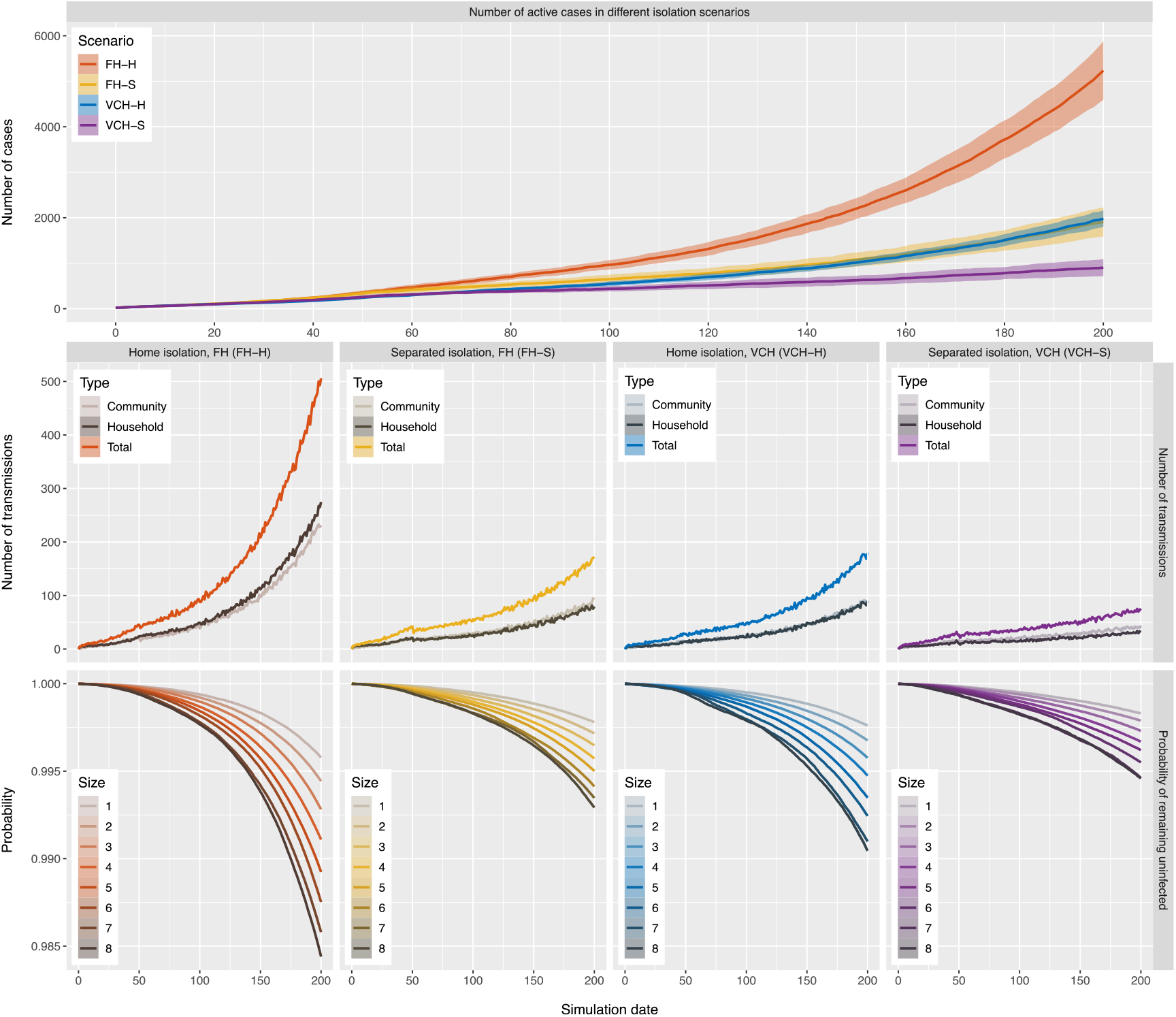
Results of simulations for isolation scenarios where 25% of the simulated individuals practice home or separated isolation. Top: the number of active cases with 95% confidence interval bands in each of the isolation scenarios. Middle: the number of transmissions of different types in each of the isolation scenarios. Bottom: the survival curves for individuals in households of different sizes in each of the isolation scenarios. All curves in the figure reflect mean values over 100 runs of the simulation for each of the isolation scenarios and the settings only differ in household size distribution and the individuals’ isolation preferences.

